# Validation of the Panbio™ COVID-19 Antigen Rapid Test (Abbott) to screen for SARS-CoV-2 infection in Sint Maarten: a diagnostic accuracy study

**DOI:** 10.1101/2021.09.23.21260526

**Authors:** Colin King, Eva Lista-de Weever, Maria Henry, Radjin Steingrover, Chérina Fleming, Richard Panneflek, Sanne van Kampen

## Abstract

**Objectives:** Control of the pandemic has required countries to look for other forms of tests besides the gold standard real-time polymerase chain reaction (RT-PCR). Rapid antigen tests (RAT), though less sensitive than RT-PCR, offer the possibility of rapid, inexpensive and early detection of the most infectious COVID-19 cases. Only very few studies have assessed the performance of the Abbott Panbio COVID-19 RAT among asymptomatic people or in Latin America. This study set out to validate this test among people attending the public test street in Sint Maarten, Dutch Caribbean.

**Methods:** People of all ages were recruited from the public COVID-19 test street regardless of COVID-19 symptoms. They received a nasopharyngeal swab for the Abbott Panbio COVID-19 RAT and the RT-PCR Qtower. Diagnostic accuracy of the RAT was compared to the RT-PCR among the overall study population and for subgroups with/without symptoms, with/without close contact and different Ct values.

**Results:** Using a RT-PCR Ct cut-off value of <33, 119 out of 1,411 people (8.4%) tested positive for SARS-CoV-2. Most were asymptomatic (59%). The overall sensitivity and specificity of the RAT was 84% (95% CI 76.2-90.1) and 99.9% (95% CI 99.6-100) respectively. The sensitivity reduced to 67.6% (95% CI: 49.5%, 82.6%) among people without symptoms, regardless of whether they were in close contact with a known COVID-19 case. Sensitivity reduced considerably with a Ct cut-off value of <35.

**Conclusions:** The Abbott Panbio RAT is a valid and cheaper alternative to RT-PCR when used on symptomatic individuals among the general population. However, among asymptomatic people it should not be used as a stand-alone test and negative results should be confirmed with RT-PCR.

## Introduction

Adequate and rapid diagnostics are essential for the treatment and control of COVID-19. ^1^ The gold standard test to detect the SARS-CoV-2 virus is real-time polymerase chain reaction (RT-PCR), a molecular assay targeting specific viral DNA.^2^ In Sint Maarten, the public health authorities use RT-PCR for SARS-CoV-2 testing to identify current infections in individuals with signs or symptoms consistent with COVID-19. This testing is also used when an asymptomatic person has been in contact with a SARS-CoV-2 positive case or identified by a positive person through contact tracing.

While RT-PCR has optimal sensitivity and specificity to detect SARS-CoV-2, it is an expensive technique that can only be performed in well-equipped laboratories with trained laboratory staff. Control of the pandemic has required countries to look for other forms of tests besides the standard RT-PCR. Rapid antigen tests (RAT), though less sensitive than RT-PCR, offer the possibility of rapid, inexpensive and early detection of the most infectious COVID-19 cases in appropriate settings.^3^ RATs generally use an immunological lateral flow assay to detect SARS-CoV-2 viral antigens.^4^ RATs would be an appropriate alternative to RT-PCR for many settings considering it would reduce the pressure on laboratories, while also expanding testing capacity. A recent study from the Netherlands and Aruba found that the Abbott Panbio COVID-19 RAT had a sensitivity of 87.4% and a specificity of 100% among symptomatic people.^5^ Only very few studies have assessed the performance of the Abbott Panbio COVID-19 RAT among asymptomatic people or in Latin America and the Carribbean.^6 7^

This study set out to compare the Abbott Panbio COVID-19 RAT to the standard molecular diagnostic assay RT-PCR among people who attended the public COVID-19 test street in Sint Maarten, Dutch Caribbean, regardless of symptoms.

## Methods

### Study population and period

The study population consisted of people from the community of Sint Maarten, of all ages, who voluntarily came to the public test street to receive a COVID-19 test. Participants were enrolled from the 11th of January 2021 until the minimal required sample size was reached. All persons were given verbal information about the testing procedure and were asked for consent to participate in the study. Each person was tested with one nasopharyngeal swab for RT-PCR and another nasopharyngeal swab for RAT by trained public health personnel. People who did not receive two nasopharyngeal swabs, who tested positive for COVID-19 in the past three months or did not consent to participate were not included in the study.

### Diagnostic tests

#### RT-PCR test

The RT-PCR samples were sent to St Maarten Laboratory Services that operates an in-house open PCR assay using a Q-tower amplification system targeting the SARS-CoV-2 E-gene. Laboratory staff was trained on this system and the system was validated by the National Institute for Public Health and the Environment of the Netherlands (RIVM). Amplification is cut off at 35 cycles.

#### Rapid Antigen Test

The RAT selected was the Panbio Abbott COVID-19 Rapid Antigen Test. This is an immunofluorescence-based lateral flow assay that can detect SARS-CoV-2 antigens within 15-30 minutes. This test fits the WHO criteria (sensitivity of ≥80%, specificity of ≥97%) ^8^ and has been validated in the Netherlands among symptomatic persons.

### Sample size calculation

Based on Buderer *et al*.,^9^ a minimal required sample size was calculated to accurately estimate the sensitivity and specificity of the RAT with a significance level of 5%. With an estimated sensitivity of 80% and specificity of 97% (WHO’s minimum requirements) and a low estimated prevalence of disease of 5%, a minimum sample size of 1,230 participants was estimated to provide a precision of 0.1 for the sensitivity’s and specificity’s 95% confidence interval.

### Analysis

Data was collected on demographics, clinical information and test results. Demographics included contact details, age, sex, occupation, and house doctor. Clinical information included COVID-19 symptoms, whether someone had been in close contact with a confirmed COVID-19 patient and the last date of contact. Test results included results from the RT-PCR and RAT tests. Participants with indeterminate or missing results were excluded from the study. The study only collected personal data that was already routinely collected when people attend the public health test street.

Participant characteristics were compared between people that tested positive and negative for COVID-19 with RT-PCR. Differences in terms of age, sex, symptoms and close contact between both groups were assessed using Pearson Chi-square tests (for proportions) and t-tests (for means) using a significance level of p=0.05.

To evaluate the accuracy of the Abbott Panbio COVID-19 RAT, RT-PCR was used as a reference test to calculate the sensitivity, specificity, positive and negative predictive values (PPV, NPV) with their confidence intervals at 95%. In this analysis, RT-PCR results were considered positive if the cycle threshold (Ct) value for amplification of the E-gene was below 33 cycles; and participants missing Ct values were removed. The clinical performance of the RAT was stratified by asymptomatic and symptomatic persons as well as by people with and without close contact to someone with COVID-19. Furthermore, a sensitivity analysis of RAT accuracy was done using various Ct cut-off values for a positive PCR result. All statistical calculations were performed in Stata and SPSS.

### Ethical approval

Ethical approval was requested from the Sint Maarten Medical Center (SMMC) Medical Ethics Committee, who decided that ethical oversight was not required for this study.

## Results

### Population characteristics

From the 11th of January 2021 to the 26th of February 2021 a total of 1,507 participants were recruited through the public health test street. Thirty participants (2%) had one or two missing test results and were excluded from analysis.

Of the 1,477 individuals included in the study, the median age was 40.5 years (SD 16), most were female (59%), asymptomatic (59%) and had had no close contact with someone with COVID-19 (60%) (Table 1). Most commonly reported symptoms were headache (17%), cough (17%), runny/stuffy nose (17%), and sore throat (15%).

**Table 1.** Population characteristics of enrolled participants by COVID-19 status.

| Variable | Number (%) <sup>1</sup> | COVID-19 RT-PCR + (%) | COVID-19 RT-PCR – (%) | X2 test (p-value) <sup>2</sup> |
| --- | --- | --- | --- | --- |
| **Total** | 1,477 (100%) | 202 (100) | 1,275 (100) | - |
| **Age in years (mean, SD)** | 40.5 (16.3) | 42.3 (16.2) | 40.2 (16.3) | 0.0912 |
| **Sex** |  |  |  |  |
| Male | 608 (41.3) | 69 (34.7) | 539 (42.3) | 0.041064 |
| Female | 864 (58.7) | 130 (65.3) | 734 (57.7) | - |
| **Symptoms** |  |  |  |  |
| No symptoms | 866 (59.2) | 65 (32.3) | 801 (63.5) | <0.00001 |
| With symptoms | 596 (40.8) | 139 (69.2) | 460 (36.5) | - |
| Fever | 160 (10.9) | 47 (23.4) | 113 (9.0) | <0.00001 |
| Cough | 242 (16.6) | 66 (32.8) | 176 (14.0) | <0.00001 |
| Runny / stuffy nose | 241 (16.5) | 51 (25.4) | 190 (15.1) | 0.000255 |
| Sore throat | 221 (15.1) | 38 (18.9) | 183 (14.5) | 0.106348 |
| Shortness of breath | 44 (3.0) | 11 (5.5) | 33 (2.6) | 0.027753 |
| Loss of taste or smell | 79 (5.4) | 45 (22.4) | 34 (2.7) | <0.00001 |
| Headache | 251 (17.2) | 50 (24.9) | 201 (15.9) | 0.001808 |
| Body ache | 132 (9.0) | 44 (21.9) | 88 (7.0) | <0.00001 |
| Nausea / vomiting | 48 (3.3) | 6 (3.0) | 42 (3.3) | 0.798434 |
| Diarrhea | 38 (2.6) | 6 (3.0) | 32 (2.5) | 0.711203 |
| Fatigue / tiredness | 99 (6.8) | 22 (10.9) | 78 (6.2) | 0.01304 |
| Other | 81 (5.5) | 15 (7.5) | 66 (5.2) | 0.199571 |
| **Close contact** |  |  |  |  |
| Yes | 517 (40.2) | 67 (38.1) | 450 (40.5) | 0.540245 |
| No | 770 (59.8) | 109 (61.9) | 661 (59.5) | - |

*Legend. RT-PCR: real-time polymerase chain reaction; X2: Chi-square; SD: standard* *deviation. <sup>1</sup> Number of missing results by variable: age (7); sex (5); symptoms (15); close* *contact (190). <sup>2</sup> For age, t-test was used.*

A total of 202 (14%) participants tested positive for SARS-CoV-2 with RT-PCR. Among them, the median age was 42.3 years (SD 16.2), most were female (65%) and had had no close contact (62%). An impressive 69% of positive participants reported symptoms. Symptoms significantly associated with testing positive for SARS-CoV-2 were fever, cough, loss of taste or smell, body ache (all p<0.00001) as well as runny/stuffy nose, headache, fatigue and shortness of breath (all p<0.05). Age and having been in close contact with a known COVID-19 case were unrelated.

### Diagnostic accuracy

For the accuracy analysis, 66 participants were excluded because they had missing RT-PCR Ct values. Of 1,411 people tested, 119 (8%) were positive by RT-PCR and 101 (7%) by RAT; 1,292 (92%) were negative by RT-PCR and 1,310 (93%) by RAT (Table 2). This resulted in an overall performance of the RAT with a sensitivity of 84.0%, specificity of 99.9%, PPV of 99.9% and NPV of 98.5% (Table 3). Among people with symptoms, 15% were positive for COVID-19 by RT-PCR and the RAT had an increased sensitivity of 91%. Among people without symptoms, only 4% had COVID-19 and the sensitivity of the RAT decreased to 67.7%. The performance of the RAT did not change when asymptomatic people had had close contact with a known COVID-19 case. All subgroups had a specificity of at least 99.7%; a NPV of at least 98%; and PPV of at least 91.7%.

**Table 2.**
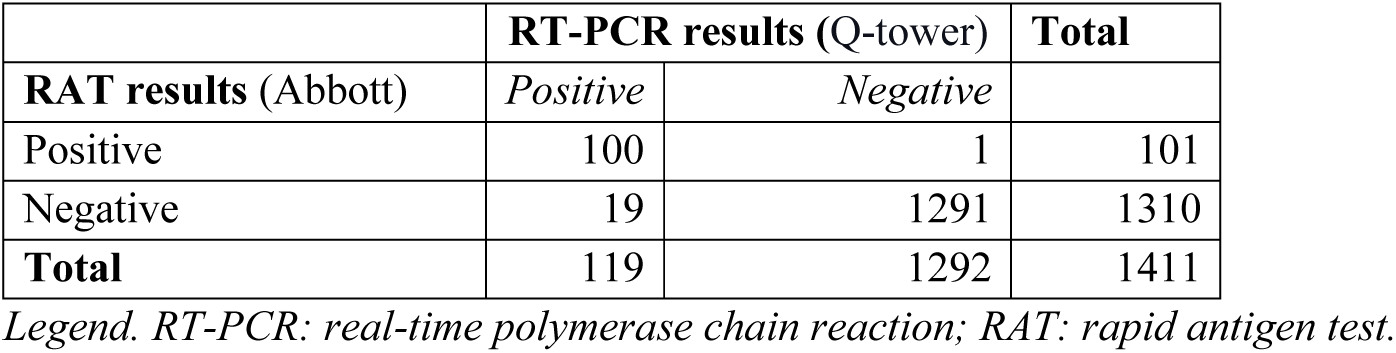
Diagnostics results among all participants (n=1,411) with PCR Ct cut-off of <33.

**Table 3.** Diagnostics results for different subgroups, with PCR positive results with a cycle threshold cut-off of <33.

| Participants | n | COVID-19<br>RT-PCR + | Prevalence<br>(95%CI) | Sensitivity<br>(95%CI) | Specificity<br>(95%CI) | PPV (95%CI) | NPV (95%CI) |
| --- | --- | --- | --- | --- | --- | --- | --- |
| All participants | 1411 | 119 | 8.4%<br>(7.0, 10.0) | 84.0%<br>(76.2, 90.1) | 99.9%<br>(99.6, 100) | 99.0%<br>(93.4, 99.9) | 98.6%<br>(97.8, 99.0) |
| With symptoms | 550 | 84 | 15.3%<br>(12.4, 18.6) | 90.5%<br>(82.1, 95.8) | 100%<br>(99.2, 100) | 100% | 98.3%<br>(96.8, 99.1) |
| Without symptoms | 846 | 34 | 4.0%<br>(2.8, 5.6) | 67.7%<br>(49.5, 82.6) | 99.9%<br>(99.3, 100) | 95.8%<br>(76.2, 99.4) | 98.7%<br>(97.8, 99.2) |
| With close contact | 370 | 17 | 4.6%<br>(2.7, 7.3) | 64.7%<br>(38.3, 85.8) | 99.7%<br>(98.4, 100) | 91.7%<br>(60.1, 98.8) | 98.3%<br>(96.9, 99.1) |
| No close contact | 374 | 13 | 3.5%<br>(1.9, 5.9) | 61.5%<br>(31.6, 86.1) | 100%<br>(99.0, 100) | 100% | 98.6%<br>(97.3, 99.3) |

*Legend. RT-PCR: real-time polymerase chain reaction; CI: confidence interval.*

### Sensitivity analyses

The diagnostic accuracy of the RAT compared to RT-PCR was analysed for Ct values of below 35, 30 and 25 (Table 4). Test sensitivity decreased as the Ct cut-off value increased. Especially when a Ct cut-off value of below 35 was used, meaning that samples with very low viral load were still considered positive, the sensitivity of the RAT decreased drastically. Specificity was at least 98.7% for all Ct cut-of values.

**Table 4.** Diagnostic accuracy of rapid antigen test compared to RT-PCR with different Ct values as cut-off for positive results.

| RT-PCR<br>positives | Number<br>(%) | Prevalence%<br>(95% CI) | Sensitivity%<br>(95% CI) | Specificity%<br>(95% CI) |
| --- | --- | --- | --- | --- |
| Ct < 35 | 136 (100 %) | 9.6% (8.1, 11.3) | 73.5% (65.3, 80.7) | 99.9% (99.6, 100) |
| Ct < 30 | 108 (79.4%) | 7.7% (6.3, 9.17) | 90.7% (83.6, 95.5) | 99.8% (99.3, 100) |
| Ct < 25 | 86 (62.2%) | 6.1% (4.9, 7.47) | 97.7% (91.9, 99.7) | 98.7% (98, 99.3) |

*Legend. . RT-PCR: real-time polymerase chain reaction; CI: confidence interval; Ct: cycle threshold.*

## Discussion

This study found that among the general public of Sint Maarten, the Abbott Panbio COVID-19 RAT identified SARS-CoV-2 infected subjects with an overall sensitivity of 84.0% and specificity of 99.9%. Most participants who attended the national test site were asymptomatic, and half of the asymptomatic people had been in close contact with a COVID-19 positive person. Compared to symptomatic people, the sensitivity was drastically lower among asymptomatic people (67.6%) and below the minimally recommended sensitivity of 80% for a RAT as recommended by WHO and far below the 90% as recommended by ECDC.^10^ The NPV among this group remained at 98.7% due to the low prevalence of COVID-19. Although there were slight variations in the specificity for different subgroups, it always stayed above the recommended 97%.

The 84% overall sensitivity of the Abbott Panbio COVID-19 RAT estimated in this study was very similar to that found in studies using the same Ct cut-off value of <33. For example, two prospective studies in community testing sites in Switzerland and Germany found similar sensitivities of 89.7% and 88.3%, respectively.^11^ Two studies in Spain that evaluated the Abbott Panbio RAT in primary health care centers reported lower overall sensitivities of 71.4% and 79.6% respectively, which may be due to the higher Ct cut-off value used.^12 13^ A real-life validation of the Abbott Panbio RAT in the Netherlands found higher sensitivities of 95.2% at the study site in Utrecht and 98% in Aruba. This was a result of testing only symptomatic people and using a lower Ct threshold for positive results. A recent study in the Netherlands focused specifically on asymptomatic football players found an overall sensitivity of 66.7%, which is very similar to the asymptomatic results from our study (67.6%).^14^ Despite the differences in sensitivity of the different studies, all had a specificity of over 99%.

One strength of our study is that it is one of the first COVID-19 diagnostic accuracy studies in the Caribbean, which provides insights into the potential performance of the Abbott Panbio RAT for other Caribbean islands. The findings can also help different islands to decide on how to best incorporate this test into their COVID-19 screening and diagnostic algorithms. Furthermore, this is one of the first studies to specifically include an analysis of asymptomatic participants and showed that it has low accuracy among this group. This may help countries to prioritize this test for symptomatic people – especially those with fever, cough, loss of taste or smell and body ache – and thereby ensure early detection and fast contact tracing of COVID-19 cases.

One limitation of this study was that it was unable to follow up persons before or after their test result. It is thus unknown whether PCR-positive persons were in the early or late stage of their infection and how the Abbott RAT performed in identifying people in their most contagious period. Further, symptoms were self-reported and may have been over-or underreported, which could have biased the difference in sensitivity between symptomatic and asymptomatic individuals.

One main implication of this study is that the Abbott RAT is not beneficial as a stand-alone test for asymptomatic people as it will miss approximately one out of three people with COVID-19 in this subgroup. The results show that approximately one out of ten positive individuals with symptoms will not be spotted with the RAT, which makes it an acceptable alternative for this subgroup. These findings confirm that COVID-19 RATs perform better among people with higher viral loads.^15^ Based on these findings, the public health authorities in Sint Maarten decided to perform the Abbott Panbio RAT among symptomatic people only, while remaining to test all people attending the test street regardless of symptoms with RT-PCR.

In conclusion, the Abbott Panbio RAT is a valid and cheaper alternative to RT-PCR for detecting COVID-19 when used on symptomatic individuals among the general population. Taking into consideration the low cost, user-friendliness, and turnaround time, this test would also be beneficial to identify clusters or outbreaks in specific settings when speed of results is of essence. However, among asymptomatic people the Abbott Panbio RAT should not be used as a stand-alone test and negative results should be confirmed with RT-PCR.

## Data Availability

The full database is available upon reasonable request from the last author (SvK).

## Acknowledgements

The authors would like to thank the staff and volunteers of the testing team at the Department of Collective Prevention Services of the Ministry of Health, Social Development and Labor of Sint Maarten for conducting data collection and field testing with the Abbott Panbio COVID-19 Rapid Antigen Test.

## Access to data

The full database is available upon reasonable request from the last author (SvK).

## Contributions

SvK, EdW, MH and RP conceptualized the study. SvK, RS and CF designed the methodology. RS and CF conducted the laboratory tests. SvK and CK supported field testing and data collection. CK conducted data validation, cleaning and analysis. CK wrote the first draft manuscript. SvK supervised all aspects of the study. All authors reviewed and edited the final manuscript.

